# Modeling the Risk of Airborne Transmission of Respiratory Viruses in Microgravity

**DOI:** 10.1101/2024.09.06.24313167

**Authors:** Chayanin Sararat, Natnicha Jiravejchakul, Kawin Nawattanapaiboon, Charin Modchang

## Abstract

Airborne transmission is the most efficient and widespread route of viral spread, posing a significant challenge in controlling major infectious diseases such as COVID-19 and influenza. In microgravity environment, such as the International Space Station (ISS), this mode of transmission requires heightened vigilance and preventive measures due to the prolonged suspension of virus-laden particles, which increases the risk of infection. Using the COVID Airborne Risk Assessment (CARA) tool, we assess the risk of airborne transmission of respiratory viruses in microgravity by simulating the emission, dispersion, and inhalation of virus-laden particles. Our findings show that the unique conditions of microgravity allow these particles to remain airborne for significantly longer periods compared to Earth, leading to a 286-fold increase in virus concentration in the air, resulting in nearly twice the probability of infection for a susceptible host. We also evaluated the effectiveness of preventive measures, and found that facemasks can reduce the risk by up to 23% while continuous HEPA filtration at five air changes per hour proves crucial for managing air quality and minimizing infection risks by reducing airborne virus concentration at 99.79%. However, when simulated the infection risk by accounting the spaceflight-induced immune suppression, we found that the infection probability increased by 12% in the condition that viral load in infected host increase for 8-fold and absence of protective measures. Although facemasks and air filtration help mitigate the risk, their effectiveness diminishes when the viral load carrying by host is high. Enhancing host immunity through vaccination or other interventions is vital, potentially reducing infection probability by up to 14.17% when combined with HEPA filtration. These findings highlight the need for robust mitigation strategies to safeguard the health of astronauts against airborne pathogens during future space missions.

## Introduction

The study of airborne transmission of respiratory viruses has gained significant attention in recent years, particularly in light of global health challenges such as the COVID-19 pandemic ^1,2^. Understanding the dynamics of how these viruses spread in various environments is crucial for developing effective prevention and control measures. One area that remains relatively underexplored is the behavior of respiratory virus transmission under microgravity conditions, such as those experienced by astronauts aboard the International Space Station (ISS). Although the likelihood of an astronaut carrying a pathogen and eventually causing an infection is small, once an infection does occur in microgravity conditions, the limited healthcare facilities on the ISS could exacerbate the situation, making it more challenging to manage and treat. Our hypothesis posits that in such conditions, respiratory particles emitted from a pathogen-carrier could remain airborne for longer durations, thereby increasing the risk of transmission among astronauts.

Spaceflight creates a uniquely stressful environment for astronauts ^3^. In addition to the effects of microgravity, astronauts encounter various stressors, including confinement in an unfamiliar environment, isolation, separation from family, sleep deprivation, noise, and anxiety. These factors have the potential to weaken the immune system during missions ^4^. Research has shown significant alterations in immune cell numbers ^5,6^, their functions ^5-7^, development ^5-7^, and cytokine secretion ^5,8^. These changes suggest that the immune system may be compromised during spaceflight, potentially increasing the host susceptibility, and reactivation of asymptomatic and latent viral infection.

One notable indicator of immune system degradation in space is the reactivation of herpesviruses ^9,10^. It is estimated that up to ninety percent of the human population, including most astronauts, is infected with herpesviruses ^11^. These viruses typically remain in a latent or dormant state after the initial infection and are usually asymptomatic in individuals with a competent immune system ^12^. However, herpesvirus reactivation has been documented even in astronauts who are in excellent health and superb physical condition ^4,9,11,13-15^. This finding highlights the compromised state of their immune systems and potentially elevates the overall risk of disease among crew members.

In the context of respiratory disease transmission, the unique microgravity environment aboard the ISS may further exacerbate the risk. Under normal gravity conditions on Earth, respiratory particles expelled by an infected individual tend to settle relatively quickly due to gravitational forces ^16,17^. However, in the microgravity environment of the ISS, these particles may remain suspended in the air for extended periods, increasing the likelihood of inhalation by other crew members. This prolonged suspension, coupled with the confined living quarters and the recirculation of air within the ISS, could create conditions conducive to the spread of respiratory viruses. Investigating the potential risks associated with this unique environment is essential for developing effective strategies to mitigate the transmission of infectious diseases during spaceflight where medical facilities are limited.

To address these concerns, it is essential to conduct a comprehensive risk assessment of airborne transmission under microgravity conditions. By understanding the potential risks astronauts face during transmission events and evaluating the effectiveness of control measures in this unique environment, we can develop targeted strategies to mitigate the spread of respiratory viruses aboard the ISS. This knowledge is not only crucial for enhancing astronaut safety but also has broader applications in understanding infectious disease transmission dynamics in confined spaces and extreme environments.

In this study, we aim to quantify the risk of airborne transmission of respiratory viruses under microgravity conditions by adapting the COVID Airborne Risk Assessment (CARA) tool ^1^. We will simulate the emission, dispersion, and inhalation of virus-laden particles in the ISS environment, taking into account the effects of microgravity on particle behavior and the potential impact of spaceflight-induced immune system changes on virus transmission and host susceptibility. By comparing the results with those obtained under normal gravity conditions, we seek to provide insights into the potentially increased risk of transmission in microgravity and propose evidence-based mitigation strategies to safeguard the health of astronauts during spaceflight. The findings of this study will contribute to the development of comprehensive infectious disease management protocols for future space missions, ensuring the well-being of crew members in the face of unique challenges posed by the spaceflight environment.

## Results

### Risk of airborne transmission under microgravity

Low-gravity environments exhibit significantly different particle behavior dynamics compared to Earth. The reduced gravitational pull allows particles to remain suspended in the air for much longer periods than they would on our planet. For example, a particle with a 3-micron diameter can theoretically remain afloat for 17.5 years (∼1.5 million hours) under a gravity of 10^−6^ g, compared to just 1.5 hours under Earth’s gravity (**Fig. 1a**). Particles as large as 10 micrometers, which normally settle within 8 minutes on Earth, can remain suspended for 15.7 years in microgravity. This prolonged suspension time enables viruses to accumulate in the environment, leading to increased viral concentrations and potentially elevating the risk of infection transmission between individuals or across spacecraft compartments.

**Fig 1:**
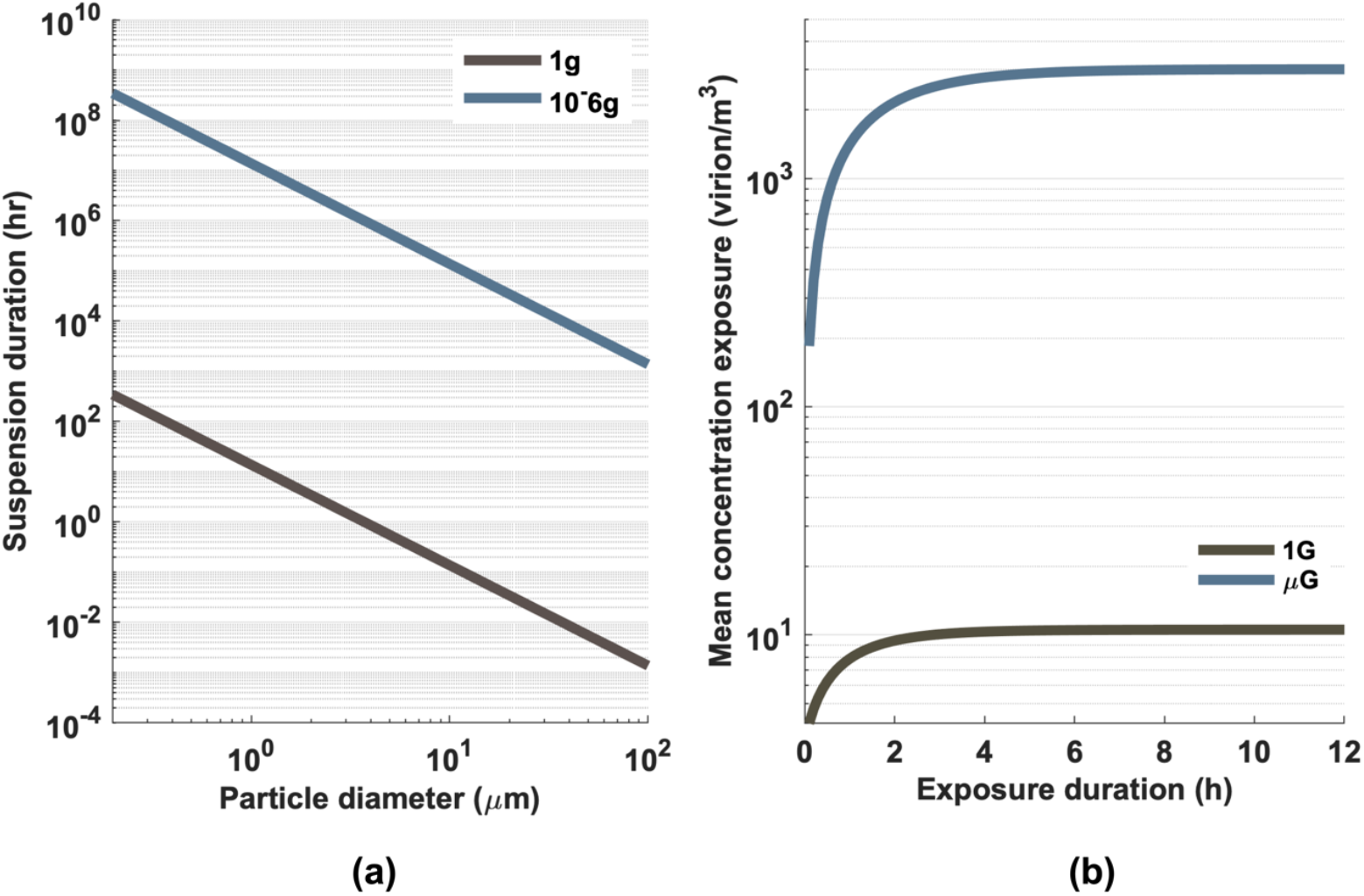
Impact of microgravity on respiratory droplet behavior and viral concentration. (a) Comparison of respiratory droplet suspension duration in Earth’s gravity (g = 9.8 m/s^2^) versus microgravity (g = 9.8 × 10^−6^ m/s^2^), illustrating the dramatically increased airborne time in reduced gravitational conditions. **(b)** Temporal evolution of SARS-CoV-2 Delta variant concentration in an enclosed space containing an infected transmitter, contrasting Earth’s gravity and microgravity environments.

In the absence of precautionary measures, the concentration of viruses in the ambient air in microgravity environments can increase by approximately 286-fold, from 10 virions/m^3^ under Earth’s gravity to 3,013 virions/m^3^ in microgravity (**Fig. 1b**). Consequently, after one week of exposure, the probability of infection for a given absorbed dose of the virus in a microgravity environment reaches 78%, which is nearly twice the infection probability under Earth’s gravity (**Fig. 2**). However, by wearing a facemask, an infectious host can reduce the number of virus-laden droplets released into the air by 84.99% (**Fig. 2a**). In this source control scenario, the infection probability decreases to 67% (**Fig. 2b**), representing a 14% reduction. Notably, this approach proves more effective than when only the susceptible host wears a mask, where the infection probability remains at 72%. The infection probability further decreases slightly to 60% when both the susceptible and infectious hosts wear masks.

**Fig 2:**
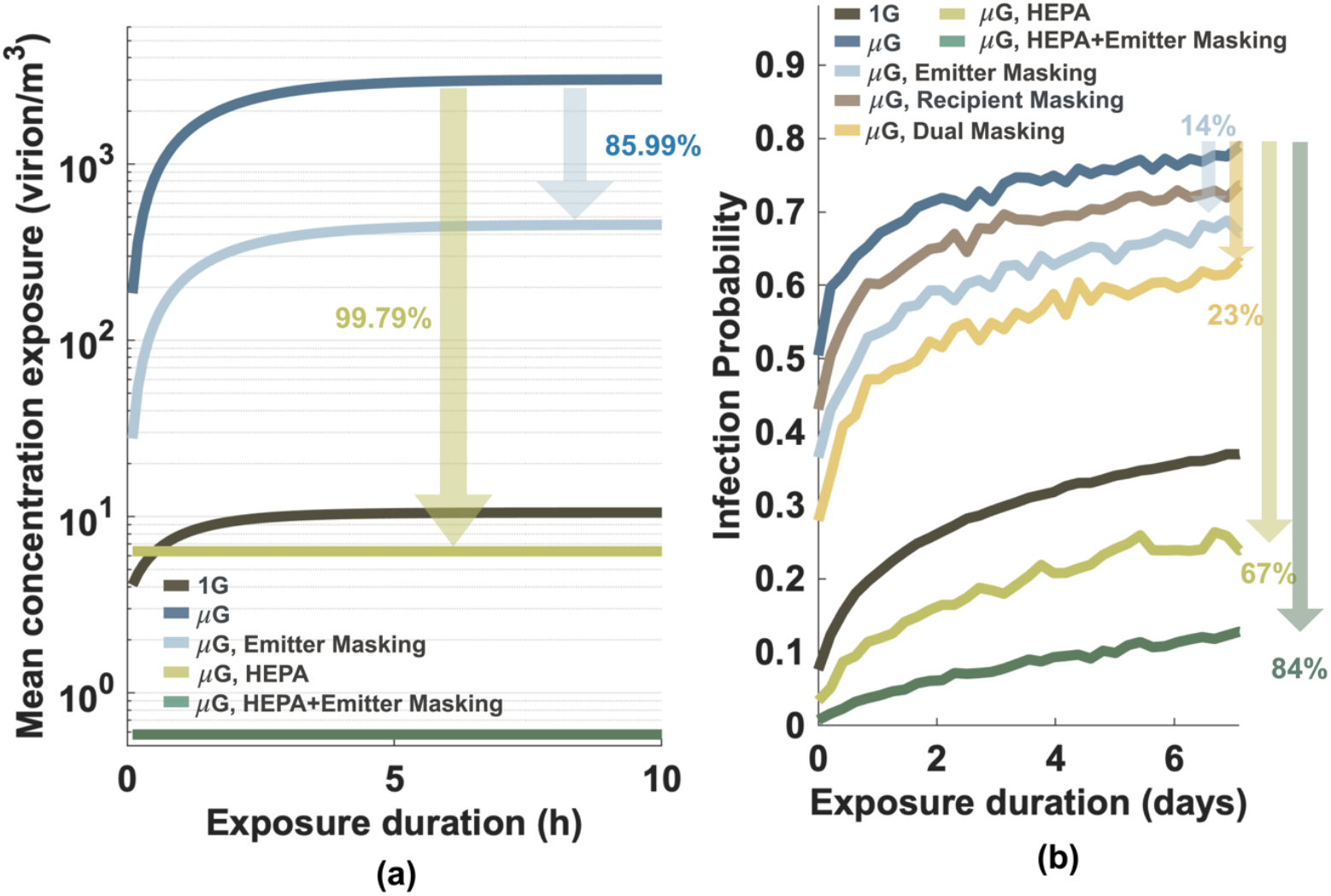
Efficacy of masking and HEPA filtration in mitigating viral transmission under microgravity conditions. (a) Reduction in airborne viral concentration and **(b)** decrease in infection probability over a 7-day exposure period under various intervention scenarios. Simulations evaluate mask-wearing by the emitter, recipient, or both, as well as HEPA filtration alone and in combination with emitter masking.

Viruses adhered to particles can be effectively removed from the air when trapped within filter media, thereby decreasing the overall viral load in the environment. High-Efficiency Particulate Air (HEPA) filters, operating continuously at 5 air changes per hour (ACH), which is comparable to standards in biological safety laboratories and hospital wards, significantly reduce the steady-state concentration of airborne viruses by 99.79%. This reduction achieves concentrations even lower than those observed under Earth’s gravity. With HEPA filters in place, the infection probability can be reduced to 25%, which is lower than the infection probability under Earth’s gravity, highlighting the importance of effective filtration systems in mitigating the risk of airborne transmission in microgravity environments.

### Effects of suppressed host immunity during spaceflight

The stress and altered conditions experienced during spaceflight can cause significant changes in immune response ^5-7^, including reduced activity of T-cells ^5,7^, which play a crucial role in fighting viral infections. Spaceflight has been shown to alter the production of cytokines ^5,18^, the signaling molecules that mediate immune responses, potentially leading to impaired immune regulation and response. Previous study has found that reactivation of herpes viruses in astronauts occurs significantly more frequently, and level of viral shedding during the spaceflight increases approximately 8-fold compared to Earth conditions ^13^. Due to the lack of data on SARS-CoV-2 shedding in space, we assumed that the viral shedding of SARS-CoV-2 would similarly increase upon the spaceflight-induced immune suppression, as seen with herpesviruses, and the enhanced shedding result in a higher viral load within the host. By assuming that the viral load in human host increased for 8 folds, our simulations showed that the infection probability is increased up to 12% in the absence of any protective measures (**Fig. 3a**). Although wearing masks can reduce the probability of infection, the risk of infection remains elevated in the condition of high viral load, even when both the transmitter and the susceptible individual are masked. Moreover, even with the implementation of a HEPA filter, the risk of infection under the 8-fold higher viral load still increases by 75%, leading to a transmission risk that is 19% higher than that observed under Earth’s gravity. Our analysis showed that microgravity could indirectly influence the transmission of SARS-CoV-2 by suppressing the host immune system, leading to the enhancement of viral shedding and increased in viral load, which further complicates the control of airborne viral transmission in spaceflight.

**Fig 3:**
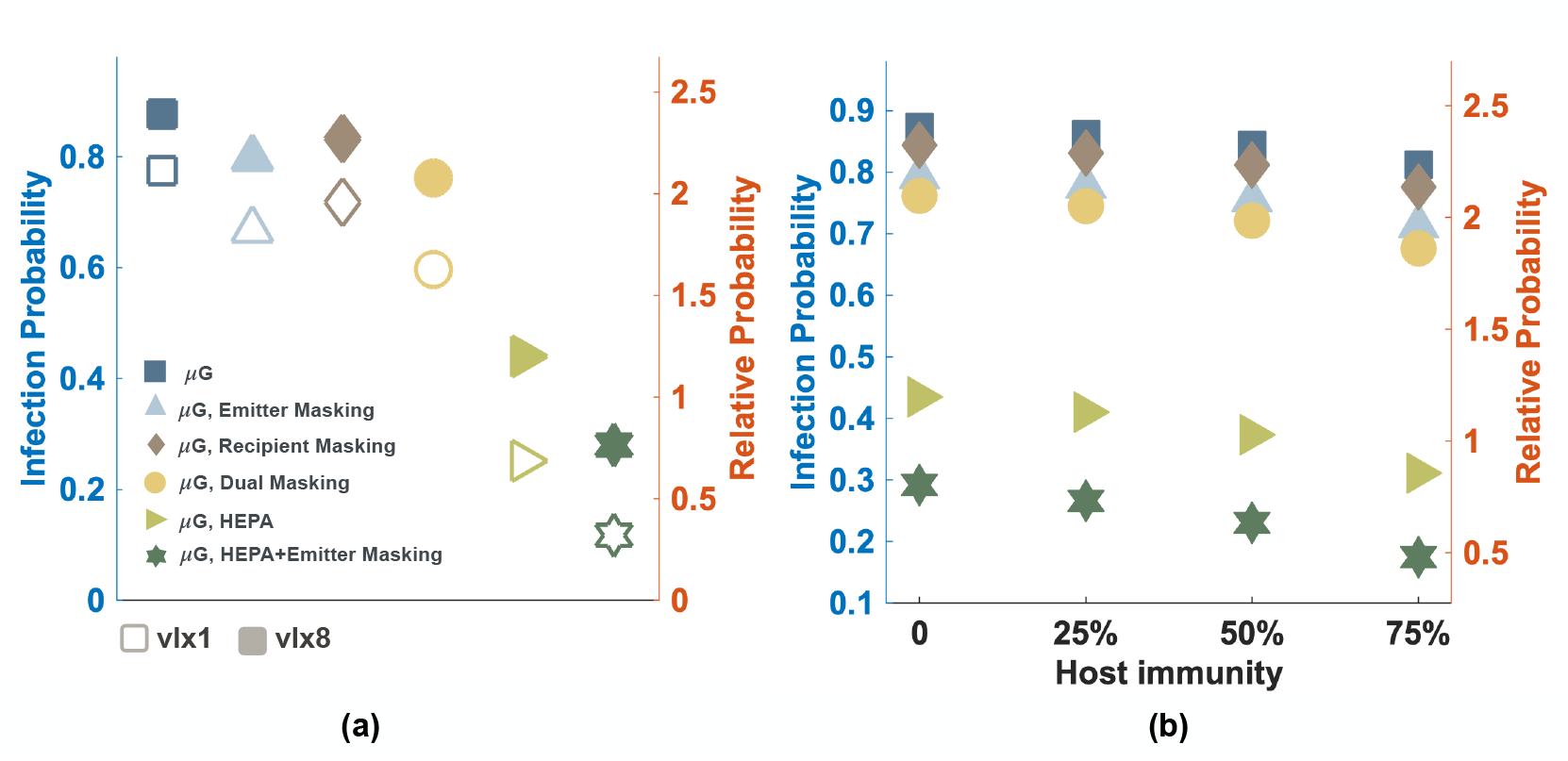
Impact of viral load and host immunity on infection probabilities in microgravity over a 7-day exposure period. (a) Comparison of infection probabilities under normal viral load versus an 8-fold increase in viral load within the infected host, illustrating the effect of spaceflight-induced immune suppression. **(b)** Infection probabilities across varying levels of immunity in the susceptible host, alongside different control measures. The left y-axis represents infection probability, and the right y-axis shows the relative probability compared to the baseline situation of transmission under Earth gravity without any measures.

Boosting host immunity through methods such as vaccination is crucial for reducing the risk of infection. Our findings demonstrate that a 50% increase in host immunity can reduce the infection probability by 3.4% without any other intervention, by 4.7% when the transmitter wears a mask, and by a substantial 14.17% when HEPA filtration is employed (**Fig. 3b**). The reduction achieved through HEPA filtration brings the infection probability down to a level comparable to our baseline scenario of transmission on Earth without any intervention. Furthermore, combining the use of a mask on the transmitter with HEPA filtration can further lower the infection probability, reducing it to a level even lower than the baseline scenario. These results highlight the importance of a multi-faceted approach to mitigating the risk of infection in the unique environment of spaceflight, where the combined effects of suppressed host immunity and prolonged particle suspension can significantly increase the likelihood of disease transmission.

## Discussion

Contamination during space flights, despite rigorous pre-launch screening, remains a possibility. Astronauts can harbor viruses and microbes in dormant or asymptomatic states, which may become active and shed later in space when the immune system is compromised. Additionally, even though astronauts are in space, the spacecraft itself constructed on Earth, and carry viruses or microbes into space, independent of the crew ^19,20^. Furthermore, if an infection does occur during space flights, the limited healthcare facilities onboard could make managing and treating the condition more challenging than on Earth.

During space flights, astronauts experience a state of microgravity where gravity-driven forces are nearly absent. Aerosols and droplets do not settle to the ground as they do on Earth. Instead, they can remain suspended in the air for extended periods. This prolonged suspension allows particles, including virus-laden droplets, to accumulate in the environment, leading to increased viral concentrations, which in turn may elevate the risk of infection transmission between individuals or across different compartments of a spacecraft. According to this modeling study, the reduced gravitational force can lead to the accumulation of viral particles in the environment and a significant increase in the concentration of airborne viruses, by approximately 286-fold. As a result, the probability of infection in a microgravity environment increases markedly due to the higher viral concentration. After one week of exposure, the infection probability can reach 78%, which is nearly double the infection probability under Earth’s gravity.

Facemasks play a crucial role in mitigating the risk of infection transmission in microgravity environments. Although combined mask usage provides the best protection, our findings suggest that preventing transmission from the source by having the infectious host wear a facemask is more effective than solely protecting the susceptible host. This implies that priority should be given to source control measures. However, it is important to note that a limitation of this work is the assumption that mask filtration performance remains consistent in microgravity as it does under Earth’s gravity. In microgravity, particle movement is governed by different forces compared to Earth, and the absence of gravitational settling alters particle behavior. On Earth, natural convection currents help move air through and around masks, assisting in the filtration process. In microgravity, these currents are greatly reduced ^21^, which might alter airflow patterns and, consequently, the mask’s filtration efficiency. Further research is needed to evaluate the performance of facemasks in microgravity conditions to ensure their effectiveness in mitigating the spread of infectious diseases.

HEPA filters are crucial for managing air quality and reducing infection risks in microgravity environments. Our findings indicate that, when operating continuously at 5 air changes per hour (ACH), comparable to standards in biological safety laboratories and hospital wards, these filters can reduce the steady-state concentration of airborne viruses by 99.79%, resulting in viral concentrations lower than those found under Earth’s gravity. On the International Space Station (ISS), high-efficiency particulate air (HEPA) filtration media are integrated within the bacteria filter elements (BFEs). The airflow through these BFEs varies between modules to maintain optimal cabin ventilation characteristics ^22^. For instance, in the U.S. Laboratory (Destiny), 6 BFEs handle an airflow of approximately 113 m^3^/hr, while in Node 1 (Unity), 4 BFEs manage about 127 m^3^/hr ^22^. Given the respective volumes of 123 m^3^ for Destiny and 80 m^3^ for Unity ^23^, this translates to air change rates of approximately 6.5 ACH and 6.4 ACH, respectively. These rates are greater than those used in our simulations, indicating a more rigorous air filtration process on the ISS, which further ensures the reduction of airborne viral concentrations and enhances overall air quality.

While astronauts are among the strongest individuals, both physically and mentally, the altered immune function during spaceflight poses significant challenges for maintaining their health. Studies have shown that reactivation of latent viruses occurs significantly more frequently during spaceflight ^11-13^, with virus shedding observed at approximately eight times the levels found on Earth ^13^. This substantial increase in viral shedding indicates that astronauts carry a much higher viral load. Although SARS-CoV-2 is not a latent virus, it can cause asymptomatic infections with an incubation period of up to 14 days. Moreover, asymptomatic carriers have also been found to shed viable viruses for up to 7 days ^24^. Apart from viruses, other airborne pathogens, such as *Mycobacterium tuberculosis*, have much longer incubation periods, ranging from a few months to up to 2 years ^25^. Both asymptomatic infections and airborne pathogens with long incubation periods may become active under the immune-suppressive conditions induced by space missions, leading to increased viral load in the infected host and greater shedding of pathogens into the enclosed air environment. Here, according to the available data on herpesviruses that viral shedding increased for 8 folds, our simulations suggest that an 8-fold increase in viral load of SARS-CoV-2 raises the infection probability by 12%. This heightened viral load poses a greater risk of transmission among crew members, even with stringent preventive measures in place. Although preventive measures such as masks and HEPA filters are typically effective in reducing infection probabilities, the higher viral loads observed during spaceflight can compromise their effectiveness. Even with HEPA filters, the infection risk increases by 75%, leading to a transmission risk that is significantly higher than on Earth. This finding highlights the need to re-evaluate and enhance current preventive strategies for space missions to account for the unique conditions and increased risks associated with spaceflight.

Boosting host immunity through methods such as vaccination plays a crucial role in reducing the risk of infection. Our results demonstrate that when immunity is boosted by more than 50%, coupled with HEPA filtration, the infection probability can be reduced to a level comparable to our baseline scenario of transmission on Earth without any intervention. This emphasizes the effectiveness of enhanced air filtration systems in mitigating infection risks, particularly when combined with strategies to improve host immunity. Moreover, employing multiple layers of preventive strategies, such as combining a mask on the transmitter with HEPA filtration, provides a compounded effect that surpasses the reduction achieved by HEPA filtration alone. This multi-faceted approach is particularly effective in high-risk environments, such as spaceflight, where maintaining low infection rates is of utmost importance. By implementing a comprehensive strategy that addresses both environmental controls and host immunity, the risk of infection transmission can be significantly reduced, ensuring the health and safety of astronauts during long-duration space missions.

In real-world settings, COVID-19 vaccines have demonstrated high effectiveness in preventing SARS-CoV-2 infection, with effectiveness ranging from 64% to 90% in the general population ^26,27^. However, in immunocompromised groups, such as individuals with certain medical conditions or those undergoing treatments that suppress the immune system, vaccine effectiveness can be lower. A study conducted in the US from March 1 to July 31, 2021, reported a vaccine effectiveness of 79% in non-immunocompromised individuals, compared to 64% in the immunocompromised population ^28^. This situation is comparable to astronauts, who often experience suppressed immune function, potentially resulting in similarly reduced vaccine effectiveness. These findings highlight the necessity of tailored preventive measures and continuous health monitoring for vulnerable populations, including astronauts, to ensure optimal protection against infections and maintain their overall health and safety.

Although radiation exposure in the space environment is higher than on Earth, the ISS is designed to shield astronauts sufficiently from this harm ^29^. As a result, the effect of radiation was not considered in our analysis. It is important to note that viruses have the potential to adapt and survive in new environments, but evidence on their effects on hosts in the context of spaceflight is not yet clear. Microgravity may alter how viruses spread, survive, and mutate. Research that reveal new insights into viral evolution, transmission patterns, and potential changes in virulence in microgravity environment is required. Research on the variability of viromes in spacecraft is limited compared to studies on bacterial and fungal microbiomes ^19^. This scarcity in research is due to factors such as the dependence of viruses on a host, their low biomass, and their complex phylogeny ^30^. However, some studies have revealed that viruses can adapt in ways that have both positive and negative effects on hosts. For example, microgravity can boost factors that inhibit Kaposi’s sarcoma-associated herpesvirus (KSHV) ^31^. On the other hand, microgravity appears to weaken the gut’s barrier, which normally helps prevent viruses from entering the body. Studies have found that in space, the gut barrier becomes more permeable, potentially allowing viruses to spread more easily ^32^. Due to the lack of data on SARS-CoV-2, our study assumes an 8-fold increase in viral load based on the data of herpesvirus shedding. However, the biology of viruses, the routes of infection and the host-pathogen interactions for each virus are different therefore, the effect of microgravity on viral shedding and spread may differ uniquely across different pathogens. Addressing these research gaps require further studies.

Conducting quantitative research on the alteration of host immunity, particularly regarding host susceptibility to infectious disease during spaceflight presents significantly challenged. However, understanding how microgravity affects immune system parameters and enhances infection risk is crucial for designing of therapeutic interventions and development of preventive measures. This knowledge could guide the development of healthcare monitoring protocols, as well as supplements or medicines to counteract microgravity-induced immune suppression, complementing existing measures like HEPA filters and facemasks

## Materials and Methods

### Infection risk assessment

The risk of airborne transmission under microgravity conditions was assessed using the COVID Airborne Risk Assessment (CARA) tool ^1^. The CARA tool simulates the spread of respiratory viruses through three main processes (**Fig 4**):

**Fig 4:**
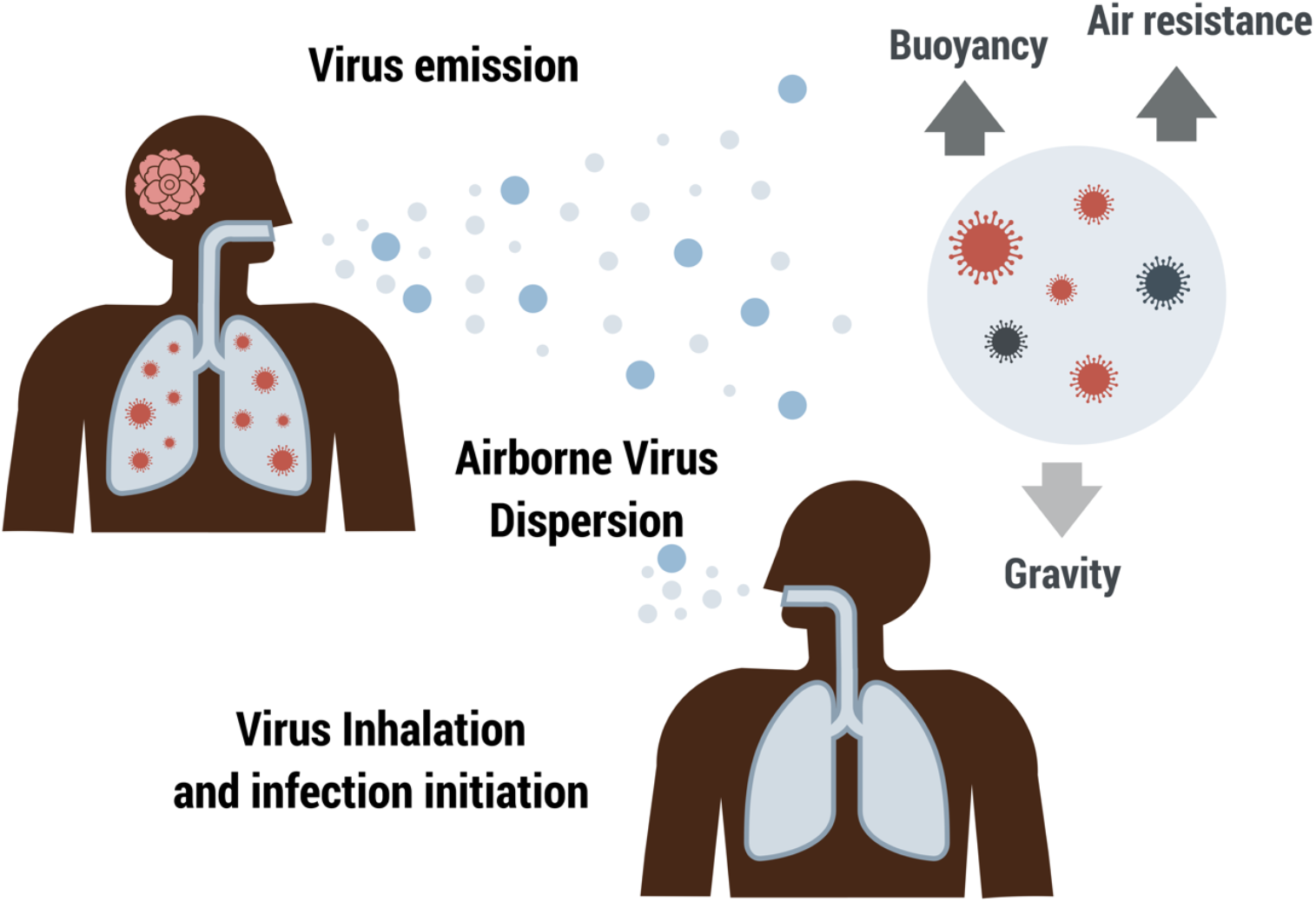
Schematic overview of the airborne transmission model for respiratory viruses in microgravity. The airborne transmission of respiratory viruses is simulated in three main stages. The first stage, *Virus Emission*, involves an infected host releasing virus-laden particles into the air through activities such as coughing, sneezing, talking, or breathing. The second stage, *Airborne Virus Dispersion*, encompasses the spread of these particles through the air. This dispersion is influenced by several factors, including gravitational settling (particles settling out of the air due to gravity), biological decay of the virus (natural degradation of the virus over time), and air management systems (e.g., ventilation and filtration systems that reduce airborne viral concentrations). The third stage, *Virus Inhalation and Infection Initiation*, occurs when a susceptible host inhales the virus-laden particles. The probability of infection is calculated based on the concentration of inhaled particles, their deposition in the respiratory tract, and the host’s immune response. The model takes into account the interplay of these processes to assess the risk of airborne transmission in a given environment.

### Virus Emission

An infected host releases virus-laden particles into the air through activities such as coughing, sneezing, talking, or breathing. The model quantifies the amount and size distribution of the emitted particles based on the type of expiratory activity and the viral load within the host.

### Airborne Virus Dispersal

Once emitted, the virus-laden particles disperse through the air. This process is influenced by several factors, including: (1) Gravitational settlement: Particles settle out of the air due to gravity, with larger particles settling faster than smaller ones. (2) Biological decay: The natural degradation of the virus over time, which depends on environmental conditions such as temperature and humidity. (3)Air management systems: Ventilation and filtration systems that reduce airborne viral concentrations by replacing contaminated air with clean air and capturing virus-laden particles.

### Virus Inhalation and Infection Initiation

A susceptible host inhales the dispersed virus-laden particles. The model calculates the probability of infection based on the concentration of inhaled particles, the deposition efficiency of particles in the respiratory tract, and the host’s immune response. The deposition efficiency varies with particle size, with smaller particles penetrating deeper into the lungs.

Detailed equations and parameters used in the model are provided in the supplementary material.

### Simulation of Microgravity Environment

In this section, we describe the environmental settings used to simulate a microgravity environment, such as that found on the International Space Station (ISS). The ISS orbits Earth in a microgravity state, where gravity is not zero but ranges from 10^−3^ to 10^−6^ g ^33,34^. This microgravity environment significantly influences the behavior of respiratory particles, as their motion is determined by the interplay of several forces, including buoyancy, air resistance, and the reduced gravitational force. By balancing these forces, the duration that respiratory particles remain suspended in the air (*t*) under microgravity conditions can be determined using the following formula:

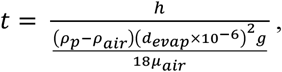

where *g* is the effective gravitational acceleration experienced in the microgravity environment, *h* represents the height from the ground to the mouth of the infected individual, *ρ*_*p*_ and *ρ*_air_ are the mass densities of the respiratory particle and air, respectively, *d*_*evap*_ is the diameter of the particle after evaporation, expressed in micrometers, and *μ*_air_ is the dynamic viscosity of air.

Variations in gravity significantly affect the time respiratory particles remain suspended in the air, which in turn influences the concentration of viruses in the air. Our study investigates gravity within the range of 10?^3^ to 10^−6^ g. However, for the specific values of 10^−4^ g, 10^−5^ g, 10^−6^ g, and 0, the results showed minimal differences (**Fig S1**). Therefore, we chose to use the 10^−6^ g value for our analysis, as it represents the most conservative estimate of the microgravity environment on the ISS.

Due to the prolonged suspension duration in microgravity, respiratory particles can remain airborne even if they are as large as 100 micrometers. Consequently, we consider respiratory particles across the entire range of inhalable sizes (1-100 micrometers ^35^) in this context, as opposed to focusing only on smaller particles that remain airborne for extended periods under Earth’s gravity.

The habitable sections of the ISS consist primarily of a series of interconnected cylindrical modules with varying volumes ^22^. These include the U.S. Laboratory (Destiny, ∼123 m^3^), European Research Laboratory (Columbus, ∼109 m^3^), Russian Service Module (Zvezda, ∼180 m^3^), Node 1 (Unity, ∼80 m^3^), Node 2 (Harmony, ∼97.3 m^3^), and Node 3 (Tranquility, ∼97 m^3^). The size of these modules directly affects the concentration of viruses within them. For this study, we simulated conditions based on the volume of the Columbus module, as it represents a mid-sized module on the ISS.

Other environmental parameters aboard the ISS are carefully controlled to ensure the health and safety of astronauts during their missions. The temperature is maintained between 20°C and 25°C, while humidity levels are controlled within a range of 30% to 60% to optimize crew performance and ensure thermal comfort ^36^. Although cosmic radiation levels are higher in space compared to Earth’s surface due to the absence of atmospheric protection, the ISS is equipped with shielding to protect astronauts from harmful radiation ^29^. Additionally, advanced air filtration and circulation systems are employed to ensure clean air for the crew, essential for maintaining a healthy breathing environment ^10^.

### Ethics declarations

This research does not involve human participants, human materials, animal subjects, or identifiable data, and we confirm that all relevant ethical guidelines have been followed in conducting the research.

## Data Availability

All data produced in the present work are contained in the manuscript

## Data availability

The authors confirm that the data supporting the findings of this study are available within the article and its supplementary information.

## Acknowledgements

This research has received funding support from the NSRF via the Program Management Unit for Human Resources & Institutional Development, Research and Innovation (PMU-B) [grant number B13F660122]. We thank Dhammika Leshan Wannigama, Mohan Amarasiri, and Somchai Chauvatcharin for their opinion on the roles of microgravity on host immunity.

## Competing interests

The authors declare no competing interests.

## Notes

### Competing Interest Statement

The authors have declared no competing interest.

