## Supplementary Material for "Modeling the Risk of Airborne Transmission of Respiratory Viruses in Microgravity"

* To whom correspondence should be addressed.

### Infection risk assessment

In this research, the risk of airborne transmission was assessed using the COVID Airborne Risk Assessment (CARA) tool [1]. The CARA tool was originally developed to evaluate the probability of being infected in a given indoor configuration. In brief, the CARA simulates the transmission process starting from the emission of virus-laden particles by an infected host, the concentration of the emitted virus in the ambient air, and then evaluates the risk of infection based on the number of viable virions inhaled by other hosts sharing the same space.

The emission rate from a source is influenced by the viral load in respiratory tract ($v$), the respiratory particle volume per unit of breathing volume ($E_p$) and the breathing rate ($B$), and can be expressed as

$$E_v = vE_pB.$$

The volume of respiratory particles emitted by an infected host per exhaled volume depends on the expiratory activity, such as breathing, speaking, or shouting, as different activities emit varying respiratory particle size profiles (**Table 1**). Additionally, physical activities influence the breathing rate of the host.

After virus-laden particles are released into the environment, these particles settle, and the pathogens can survive outside the host body for only a limited period. Additionally, viruses can be removed through air management systems. Therefore, the concentration of viruses in well-mixed room air depends not only on their emission but also on these removal mechanisms. The rate of change of the concentration of viruses ($C$) is derived from the following differential equation:

$$\frac{\partial C}{\partial t} = \frac{E_v N_I}{V} - \lambda C.$$

The first term in the differential equation represents the viral production term, while the second term represents the removal term of the virus. Here, $E_v$ is the aforementioned emission rate, $N_I$ is the number of infected hosts, and $V$ is the room volume.

The viral removal rate (λ) accounts for removal mechanisms including gravitational settlement ($\lambda_G$), biological decay ($\lambda_v$), ventilation ($\lambda_{ACH}$), and particulate filtration ($\lambda_{HEPA}$). Thus, the total removal rate is given by:

$$\lambda = \lambda_G + \lambda_v + \lambda_{ACH} + \lambda_{HEPA}.$$

The calculation of each term of the viral removal rate is summarized in **Table 2**.

Once another person enters the same space and breathes in the virus-laden particles, the probability of a COVID-19 infection ($P$) is assessed based on the intake dose of the infectious agent reaching the target infection site ($D_v$):

$$P = 1 - e^{-\frac{D_v\,(1-M)}{ID_{50}/ln2}}.$$

Even after the pathogen successfully reaches the infection site, it must survive the immune defenses to cause an infection. $M$ is the fraction of pathogens eliminated by the immune system, so $1-M$ represents the surviving fraction. The infectious dose $ID_{50}$ indicates the dose at which 50% of the population is expected to become infected. It is assumed that an infection can be initiated by a single pathogen reaching the infection site, but if the intake dose equals $ID_{50}$, the probability of infection is 50%. The calculation of infection probability and associated parameters are summarized in **Table 3**.

**Table 1: Parameters used in the viral emission rate calculation.**

| Parameter | Values | Source |
|---|---|---|
| Viral load ($v$) | Guassian kernel density with mean and sd of 6.6 and 1.7 $\log_{10}$ copies per ml | [2] |
| Volume of respiratory particles emitted by an infected host per an exhale volume for a given diameter ($E_p(D)$) | $E_p(D) = N_p(D)V_p(D)(1-\eta_{out}),$ | |
| Number of of particles of this size ($N_p$) | $N_p(D) = \frac{1}{D}\sum_{i\in\{B,L,O\}}\left[\frac{c_i f_{amp,i}}{\sqrt{2\pi}\,\sigma_i}\exp\left(-\frac{(\ln D - \mu_i)^2}{2(\sigma_i)^2}\right)\right]$<br>(Fig S2) | |
| The mean of the natural logarithm of the diameter ($\mu_i$)<br>B-mode | 0.99 (ln μm)<br>1.39 (ln μm) | [1]<br>Particle diameter |

| | | |
|---|---|---|
| L-mode<br>O mode | 4.96 (ln μm) | distribution parameters were adapted from [3], using an evaporation factor of 0.3 instead of 0.5 specified in the original source. |
| The standard deviation of the natural logarithm of the diameter ($\sigma_i$)<br>B-mode<br>L-mode<br>O mode | 0.26 (ln μm)<br>0.51 (ln μm)<br>0.59 (ln μm) | |
| Total particle emission concentrations ($c_i$)<br>B-mode<br>L-mode<br>O mode | 0.06 ($cm^{-3}$)<br>0.2 ($cm^{-3}$)<br>0.001 ($cm^{-3}$) | [4] |
| Amplification factor ($f_{amp,i}$), $i \in (B, L, O)$<br>Breathing<br>Speaking<br>Singing and shouting | (1, 0, 0)<br>(1, 1, 1)<br>(1, 5, 5) | [5] |
| Volume of each individual particle ($V_p$) | $V_p = \frac{4}{3}\pi\left(\frac{d}{2}\right)^3$ | Assumed that respiratory particles are spherical in shape. |
| Outward mask efficiency ($\eta_{out}$) | | |
| Breathing rate ($B$)<br>Seated<br>Standing<br>Light activity | Lognormal($0.51, 0.043^2$)<br>Lognormal($0.57, 0.043^2$)<br>Lognormal($1.24, 0.12^2$) | EPA Exposure Factors Handbook [6], data from [7] |

| | |
|---|---|
| Moderate activity<br>Heavy activity | Lognormal(1.77,$0.34^2$)<br>Lognormal(3.28,$0.72^2$) |

**Table 2: Calculation and parameter for the viral removal rate.**

| Parameter | Values | Source |
|---|---|---|
| **Gravitational settlement:** $\lambda_G = \frac{v}{h}$ | | |
| Approximated height from ground to mouth ($h$) | 1.5 m | |
| Settling velocity $$v = \frac{(\rho_p - \rho_{air})(d_{evap} \times 10^{-6})^2 g_E}{18\mu_{air}}$$ | Derived from Stokes' Law, by balancing the gravitational force with the drag and buoyancy forces. | |
| Mass density of the particle ($\rho_p$) | 1000 kg $m^{-3}$ | [3] |
| Mass density of the air ($\rho_{air}$) | 1.2 kg $m^{-3}$ | Calculated using the ideal gas law at normal temperature and pressure (NTP) of 20°C and 101.325 kPa |
| Diameter of the particle following evaporation ($d_{evap}$), in µm | $d_{evap} = d\, f_{evap}$ | |
| Evapolation factor ($f_{evap}$) | 0.3 | [8] |
| Gravitational acceleration of the Earth ($g_E$) | 9.81 $ms^{-2}$ | Approximated value at the surface of the Earth |
| Dynamic viscosity of air ($\mu_{air}$) | $1.8\times10^{-5}$ $kgm^{-1}s^{-1}$ | at NTP |
| **Biological decay:** $\lambda_G = \frac{\ln(2)}{t_{1/2}}$ | | |

| | | |
|---|---|---|
| Half life of a virus<br>- SARS-CoV-2, RH < 40%<br>- SARS-CoV-2, RH > 40% | 1.1 h<br>6.43 h | [9]<br>[10] |
| **Ventilation:** $\lambda_{ACH} = \frac{Q_{ACH}}{V}$ | | |
| Volumetric flow rate of fresh air supplied to a room ($Q_{ACH}$) | Depend on the type of ventilation system used, measured in $m^3h^{-1}$ | |
| Room volume ($V$) | Depend on the type of room, measured in $m^3$ | |
| **Air filtration:** $\lambda_{HEPA} = \frac{Q_{HEPA}}{V}\varepsilon$ | $\lambda_{HEPA}(0.8) = 5\ h^{-1}$,<br>comparable to the design standards for biological safety laboratories and hospital wards | |
| Volumetric flow rate of the air through the filter ($Q_{HEPA}$) | | |
| Efficiency of the filter ($\varepsilon$) | | |

**Table 3: Parameters used in the infection probability.**

| Parameter | Values | Source |
|---|---|---|
| Infectious dose required to cause infection in 50% of an exposed population (ID50) | U[10 100] | |
| Host immunity of the exposed individual ($M_{exp}$) | [0 1] | |
| Intake dose: $D_v = \int_0^{d_{max}} \int_{t_i}^{t_f} f_{inf}\, f_{dep}(d)\, (1-\eta_{in})\, B\, C(t,d)\ dt\ dd$ | | |
| Breathing flow rate (B) | See Table 1 | |
| Fraction of infectious virus ($f_{inf}$)<br>$f_{inf} = r_{inf}(1 - M_{inf})$ | | |
| Viable-to-RNA virus ratio ($r_{inf}$) | U[0.01 0.60] | |
| Host immunity of the infected individual ($M_{inf}$) | [0 1] | |

| | |
|---|---|
| Deposition fraction in the respiratory tract ($f_{dep}$)<br>$$f_{dep}(d) = I_{frac}(d)\left(0.0587 + \frac{0.911}{1+e^{4.77+1.485\ln d_{evap}}} + \frac{0.943}{1+e^{0.508-2.58\ln d_{evap}}}\right)$$<br>$$I_{frac}(d) = 1 - 0.5\left(1 - \frac{1}{1+0.00076\, d_{evap}^{2.8}}\right)$$<br>$$d_{evap} = f_{evap} d$$ | |
| Inward mask efficiency ($\eta_{in}$) | |
| Surgical mask | [0.25 0.80] |
| PPE | [0.83 0.91] |

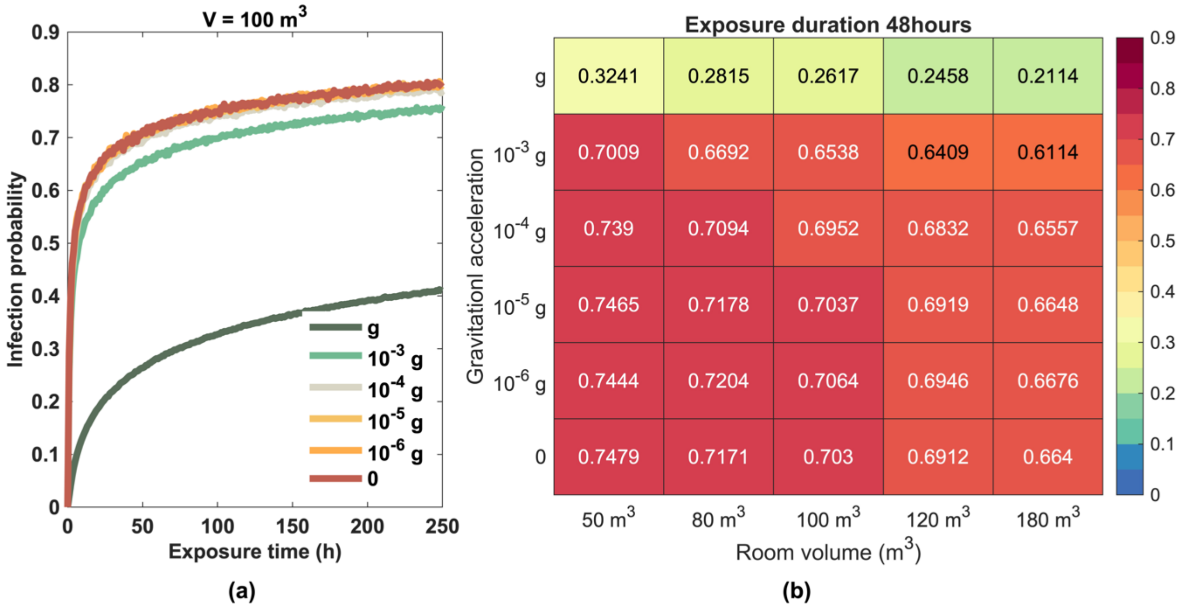

**Fig S1: Infection probability under microgravity conditions. (a)** Infection probability across various gravity levels as a function of exposure time. **(b)** Infection probability across different gravity levels and room volumes after 48 hours of exposure.
